# A Heterozygous 9q34 deletion encompassing SPTAN1 as a cause of distal myopathy

**DOI:** 10.1101/2025.01.09.24319154

**Authors:** Liedewei Van de Vondel, Jonathan De Winter, Alice Monticelli, Natacha Camacho, Tine Deconinck, Katrien Janssens, Goedele Malfroid, Alicia Alonso-Jiménez, German Demidov, Steven Laurie, Willem De Ridder, Biljana Ermanoska, Vincent Timmerman, Jonathan Baets

## Abstract

We report a family affected with childhood onset distal muscle weakness with a heterozygous chromosome 9q34 deletion encompassing the *SPTAN1* gene. The deletion was detected through exome-sequencing based copy number variant detection, segregates in four patients and is non-penetrant in two other relatives. Electromyography, muscle MRI and muscle biopsy revealed a myopathic disease phenotype. Cellular consequences of the deletion were investigated using qPCR and western blotting on patient-derived fibroblasts, which revealed a reduction of RNA but not protein levels. Immunocytochemistry was performed on muscle tissue which did not reveal reduction of α-II-spectrin. *SPTAN1* loss-of-function variants have previously been reported to cause distal hereditary motor neuropathy and recently distal myopathy. Here, we confirm the role of *SPTAN1* haploinsufficiency as a cause of distal myopathy. We propose an age-dependent lack of α-II-spectrin and suggest CNV detection in repurposed exome sequencing as an important diagnostic tool.

## Introduction

While 80% of Rare Diseases (RD) are believed to be genetic, currently only 50-60% of RD patients receive a genetic diagnosis(1,2). Detection of Single Nucleotide Variants (SNVs) and short insertions and deletions (indels) in Exome Sequencing (ES) data is standardized, while detection of larger variants (e.g. tandem repeats, copy number variants (CNVs) and structural variants (SVs)) is technically limited in short-read data. Microarrays can be used to evaluate CNVs but are seldomly performed for patients with late-onset disease. These shortcomings can be addressed by performing either (long-read) genome sequencing or by applying novel algorithms to existing ES data. Within the Solve-RD consortium(3), a large-scale effort was performed to detect CNVs in ES, using three different algorithms (ClinCNV, ExomeDepth and Conifer) and a pre-defined list containing 615 genes associated with neuromuscular diseases(4).

Heterozygous loss-of-function variants in the *SPTAN1* gene are associated with different disorders, including intellectual disability with or without peripheral neuropathy, hereditary motor neuropathy (HMN) with a variable severity and reduced penetrance, and recently also distal myopathy with variable severity(5–7). *SPTAN1* encodes α-II-spectrin, which exhibits widespread expression with an enrichment in neuronal tissues. In mice, homozygous knock-out of *Sptan1*^−/−^ is embryonically lethal, while heterozygous *Sptan1*^+/−^ mice display no phenotype(8). Neuronal-specific knock-down of *Sptan1* is critical in upholding neuronal structure(9,10). α-II-spectrin oligomerizes with one of four different β-spectrins into heterotetramers, which together with actin, form a submembrane periodic cytoskeleton which is thought to infer mechanical resilience to axons through its ability to stretch and withstand cellular forces(11).

In this study, we describe the detection and interpretation of a heterozygous 9q34 deletion encompassing the *SPTAN1* and *DYNC2I2* genes and part of the *GLE1* gene, in a multi-generational family with a childhood onset distal weakness.

## Materials and methods

### Genetic and clinical studies

Experiments were approved by the University of Antwerp ethical committee. Known genetic causes for peripheral neuropathies were excluded in participants III:2 and IV:2 by an ES-based gene panel supplemented with Human Phenotype Ontology (HPO) based analysis. ES of III:2 was submitted to the Solve-RD consortium and analyzed within the European Reference Network for Rare Neuromuscular Disorders (ERN EURO-NMD)(12,13). The identified CNV passed the filtering strategy and was reported for interpretation(4). Segregation analysis was performed using SNP array (HumanCytoSNP-12 v2.1 BeadChip Kit, Illumina) with CNV Webstore 2.0 analysis, using DNA extracted from blood in 11 family members(14). Clinical details were collected retrospectively for deceased individuals (I:1, I:2, II:1, II:2, II:3, III:3, IV:1). Individuals III:1, III:2 and IV:2 were clinically re-evaluated including Nerve Conduction Studies (NCS) and electromyography (EMG), muscle magnetic resonance imaging (MRI) was performed for patients III:2 and IV:2. Muscle biopsies were obtained respectively from the vastus lateralis (III:2) and the tibialis anterior (IV:2), and compared to a vastus lateralis biopsy from a control individual.

### Patient Fibroblasts, RT-qPCR, Western Blotting

Fibroblasts were established from III:2 and an age-and-gender matched control and cultured in Dulbecco’s modified Eagle medium (DMEM) (ThermoFisher) with 10% Fetal Bovine Serum (FBS), 1% L-glutamine and 1% penicillin/streptomycin. RNA was extracted using the Universal RNA kit (Roboklon). cDNA was generated using the High-Capacity cDNA Reverse Transcription Kit (ThermoFisher). RT-qPCR was performed using the SYBR Green PCR Master Mix (ThermoFisher), *SPTAN1* and *GAPDH* primers are available upon request. For western blotting, cell pellets were lysed with RIPA buffer and processed using the same protocol previously described(6). Primary antibodies were anti-SPTAN1 (Abcam ab11755, 1:1000), anti-GLE1 (ThermoFisher 26466-1-AP, 1:1000), anti-GAPDH (Genetex GTX100118, 1:10 000).

### Muscle Biopsy Immunocytochemistry

Seven micron cryosections of muscle tissue were fixated using ice-cold acetone and blocked using donkey serum (1:500) in PBS, and incubated overnight at 4°C with primary antibodies (anti-SPTAN1 (1:50, Abcam ab11755), anti-SPTB (1:500, Novocastra RBC2/3D5) and anti-SPTBN1 (ThermoFisher PA5-44905)). Secondary antibodies were incubated for 1 hour at 4°C, followed by autofluorescence blocking with 0,1% Sudan black for 10 minutes. Hoechst (1:20000) and phalloidin staining (1:25, Biotin) were performed. Images were acquired with a Zeiss LSM700 confocal microscope using a 20x/0.8 Plan Apochromat objective. Image analyses were performed in FIJI. Cells were segmented and masked using the Phalloidin signal as a region of interest to measure fluorescence intensity.

### Statistical Analysis

Statistical analysis was performed using R. Values were normalized to the average of control replicates. The qPCR data was analyzed using the Pfaffl method and a two-sided Student’s *t*-test. For analysis of ICC experiments, a mixed linear model was first tested, but the variance between slides was neglectable, after which a One-Way Anova was performed.

## Results

The CNV analysis identified a heterozygous deletion at locus 9q34 (figure 1A) in III:2, encompassing *SPTAN1* and was therefore reported. The deletion was called by ClinCNV and ExomeDepth and visually inspected in IGV. Segregation analysis revealed that although all affected individuals carried the variant, non-penetrance was present in two individuals (pedigree available upon request). The SNP array located the left breakpoint within *GLE1* and the right breakpoint to the intergenic region between *DYNC2I2* and *SET* (figure 1). The deletion was reported as a Variant of Unknown Significance (VUS) according to the ACMG guidelines(15).

**Figure 1:**
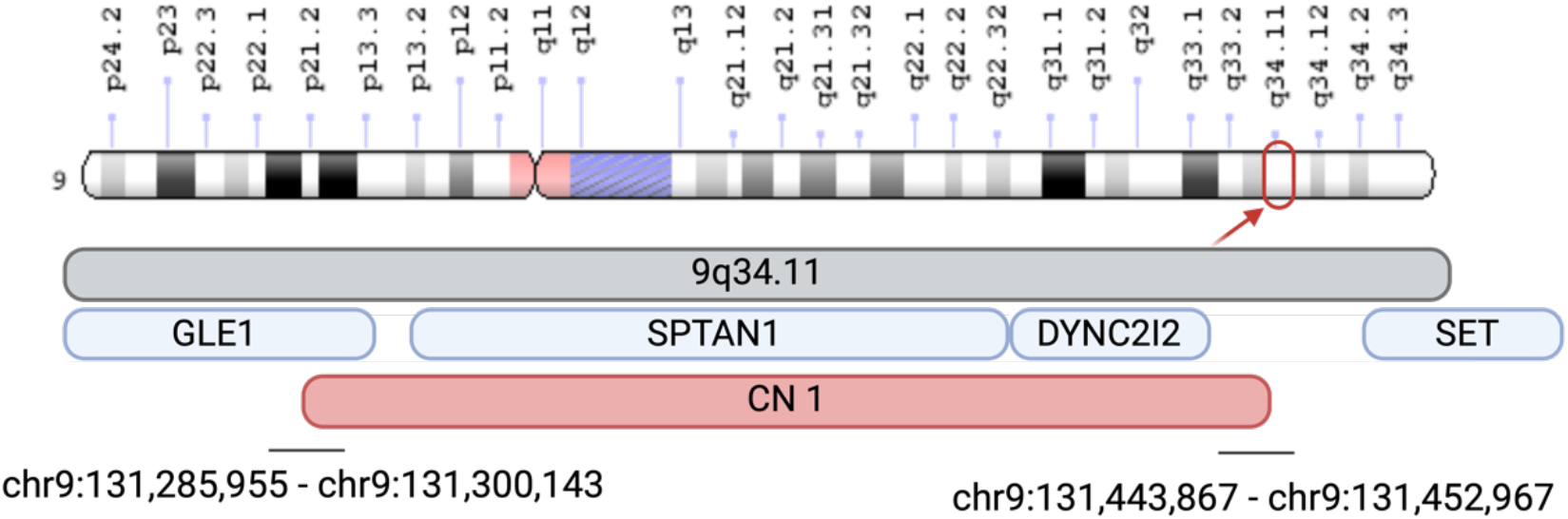
Depiction of the CNV deletion size and location. Indication of de location of the identified Copy Number Variant (CNV) in the 9q34 region with coordinates in GRCh37, including part of GLE1, SPTAN1 and DYNC2I2.

Patients exhibit varying degrees of early childhood onset lower limb distal weakness and foot abnormalities without marked disease progression or anticipation. Muscle weakness predominantly affected the anterior compartment of the distal lower limbs, selectively involved the *extensor hallucis longus* (EHL) muscle in one patient and foot dorsiflexion weakness in all affected participants. None showed evidence of proximal or bulbar muscle weakness. The extent of foot abnormalities ranged from *pes cavus* and hammertoes to distal arthrogryposis. None showed evidence of intellectual disability or facial dysmorphism. NCS and needle-EMG were inconclusive in three patients. Muscle MRI showed fatty infiltration of anterior and posterior compartments of the lower limbs in patient III:2 and bilateral absence of the EHL muscle in patient IV:2 (figure 2A and B). Muscle biopsy in III:2 and IV:2 showed myopathic changes, comprising a markedly increased number of internalized nuclei, increased fiber size variation with frequent fiber splitting and marked muscle fiber hypertrophy for participant IV:2 (figure 2C and D). Of note, patients II:3 and III:1 had a normal clinical and electrophysiological exam at ages 50-75 and are thus considered unaffected.

**Figure 2:**
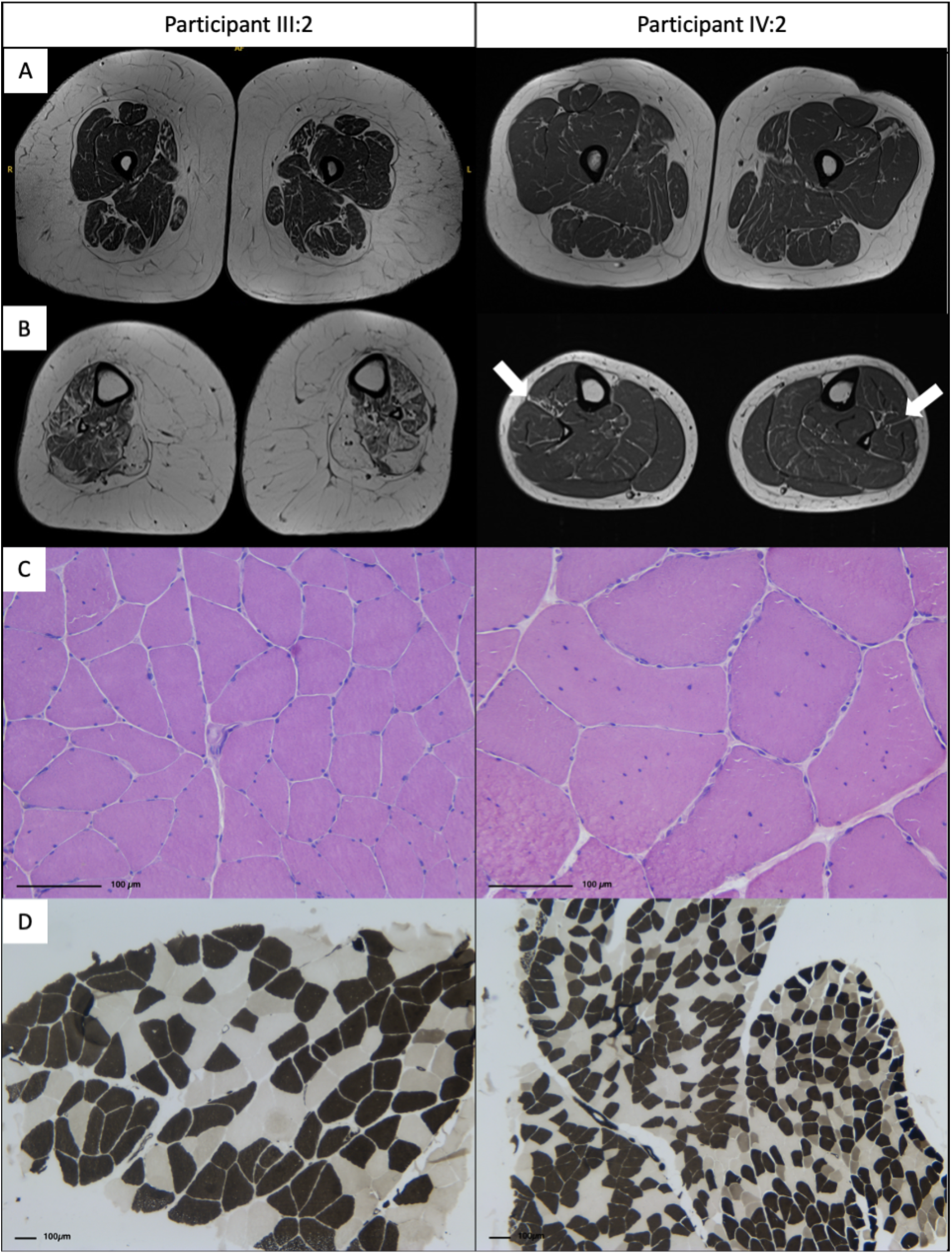
Muscle MRI and muscle biopsy findings in patients III:2 and IV:2. (A) Axial T1-weighted images at the level of the thighs. (B) Axial T1-weighted images at the level of the calfs. Patient III:2 showing diffuse fatty infiltration and muscle atrophy of both the anterior and posterior compartment. Patient IV:2 showing bilateral absence of the EHL muscles indicated by white arrows. (C) Hematoxylin and eosin (H&E) staining on muscle biopsy (III:2 right m. vastus lateralis quadriceps femoris; IV:2 right m. tibialis anterior). Myopathic features seen in both participants with increased number of internalized nuclei seen in IV:2. (D) ATPase staining (pH 4,6) showing no fiber type disproportion or atrophy.

*SPTAN1* is a known dosage-sensitive gene and in the GnomAD v4 SV database small intronic but no exonic deletions in *SPTAN1* occur (16). We hypothesized that the 9q34 deletion would lead to reduced levels of α-II-spectrin. Contrary to our assumption, α-II-spectrin levels in fibroblasts of III:2 are not reduced compared to control (figure 3A), but mRNA levels are (p = 0.0014, figure 3B). Muscle biopsies similarly did not reveal a change in the intensity of the diffuse intracellular α-II-spectrin signal measured within the phalloidin-labeled muscle fibers (figure 3C-D). Neither were protein levels of β-I-spectrin reduced, a staining commonly used to assess muscle membrane integrity and binding partner of α-II-spectrin (Supplemental figure 1). The genes adjacent to *SPTAN1* and partially deleted in this variant, *DYNC2I2* and *GLE*, are highly unlikely to be dosage-sensitive as both have a probability of being loss-of-function intolerant (pLi) score of 0(17). Furthermore, more than 2000 exonic heterozygous deletions in *DYNC2I2* are found in GnomAD v4. Recessive variants in *GLE1* cause arthrogryposis multiplex congenita,(18) and while a heterozygous loss-of-function variant was associated with Amyotrophic Lateral Sclerosis (ALS) in a single patient, this association has not been confirmed(19). The partial C-terminal deletion of *GLE1* in this family minimally includes exons 12-16 and maximally extends to exon 7. Western blot analysis on pellets from patient-derived lymphoblasts showed no evidence of a residual shorter fragment (data not shown), indicating that the deletion likely triggers nonsense-mediated decay. As *GLE1* is likely not a Mendelian dosage-sensitive gene, the patients do not exhibit clinical signs of ALS, and the pedigree suggests a dominant inheritance pattern, the deletion of the *GLE1* 3’ exons is unlikely to contribute to the disease.

**Figure 3:**
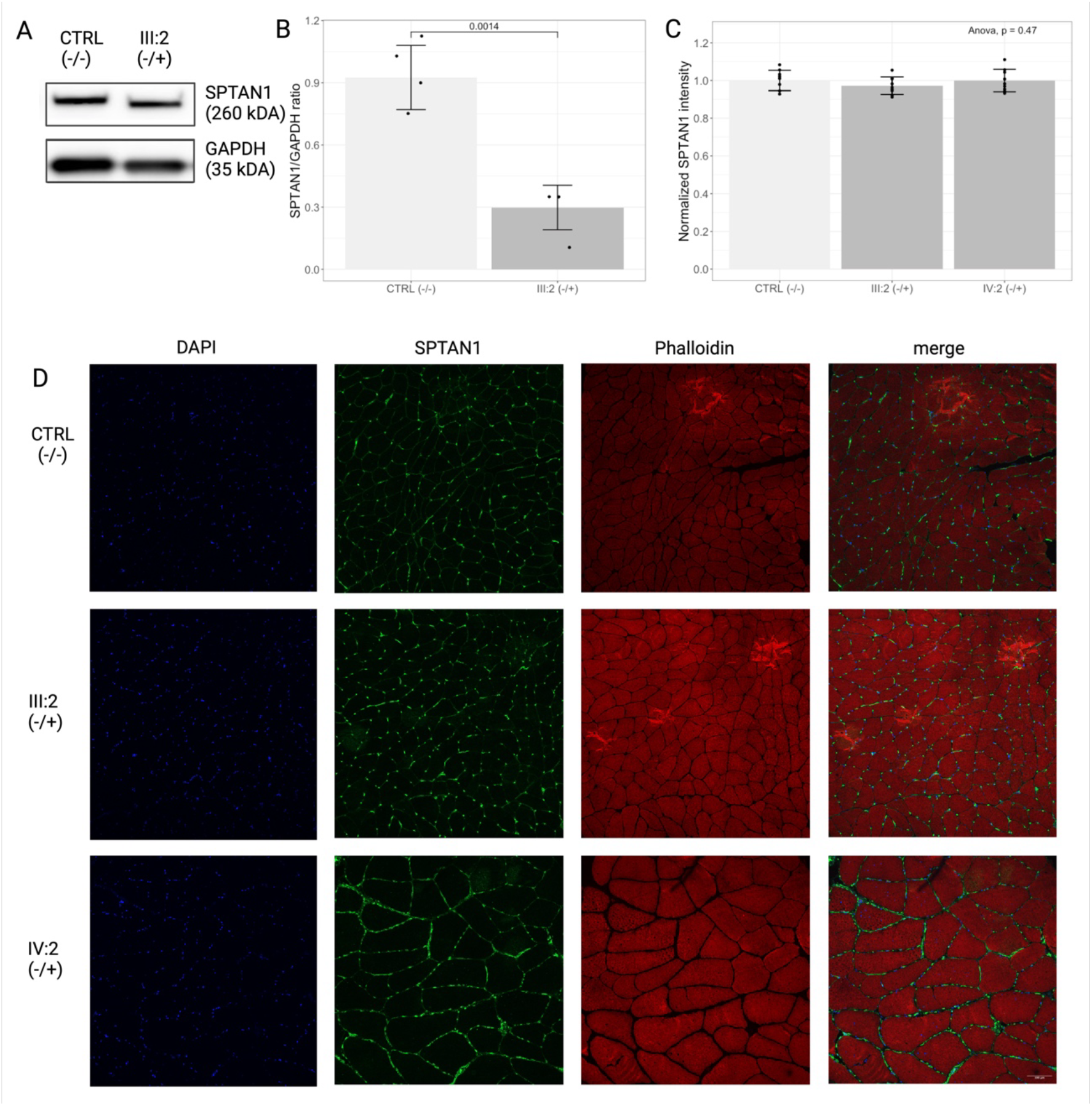
Protein, RNA and ICC measurements in patient fibroblasts and muscle biopsy. (A) representative western blot of patient-derived fibroblasts compared to control. (B) qPCR data of patient-derived fibroblasts showing a reduction of SPTAN1 mRNA. (C) SPTAN1 intensity in ICC of muscle biopsies. (D) Representative ICC images of muscle biopsies, stained for DAPI (blue), SPTAN1 (green), Phalloidin (red) and a merge image.

## Discussion

We identified a heterozygous 9q34 deletion in a family with distal myopathy through ES-based CNV analysis, warranting the addition of ES-based CNV screening to routine diagnostic practice. We propose that the heterozygous deletion of *SPTAN1* underlies the observed myopathic abnormalities in the assessed family. *SPTAN1* loss-of-function variants are known to cause HMN with reduced penetrance and have recently been reported to also cause childhood onset distal myopathy (7). Reduced penetrance and phenotypic variability are similarly observed here. This family closely resembles the reported *SPTAN1* myopathy cohort with non-to slowly progressive distal weakness and feet abnormalities with onset in early childhood.

Heterozygous deletion of *SPTAN1* reduces mRNA levels in fibroblasts, but surprisingly, protein levels are unaffected in both fibroblasts and muscle. Protein and RNA levels were equally not significantly reduced in muscle tissue of loss-of-function carriers in a recently reported study (7). Correspondingly, a conditional knock-out of mouse *Sptan1* under a myogenin driver does not result in a muscle phenotype in animals aged to six months(20), although it is known that expression of α-II-spectrin in rat skeletal muscle cells have a changing expression pattern during development (21). In *Drosophila*, the spectrin complex was shown to have a role in the fusion of myoblast precursors into multi-nucleated myofibers (22). We hypothesize that an age-related haploinsufficiency of the spectrin complex might affect this role, thereby causing a developmental muscle phenotype. A further possibility is that the spectrin complex, known to act as a molecular scaffold in many cell types, has a role in upholding the nuclear localization in the myofiber, reflected in the internalized nuclei on the patients’ muscle biopsy. Further research into the developmental expression and muscle function of the spectrin complex will prove useful to gain insight into these possible mechanisms.

## Data Availability

All data produced in the present study are available upon reasonable request to the authors.

## Acknowledgments & Funding

Several authors are member of the European Reference Network for Rare Neuromuscular Diseases (ERN EURO-NMD, project N°870177). We thank the participating patients and relatives for their cooperation in this study. J.B. and V.T. are part of the μNEURO Research Centre of Excellence of the University of Antwerp. This work was supported by the EU Horizon 2020 program (Solve-RD under grant agreement N°779257). J.B. is supported by a Senior Clinical Researcher Mandate of the Research Fund – Flanders (FWO) under grant agreement N°1805021N. L.V.d.V. is supported by a predoctoral fellowship of the FWO under grant agreement N°11F0921N. J.D.W. is supported by the Goldwasser-Emsens fellowship.

---

**Supplementary Figure 1:**
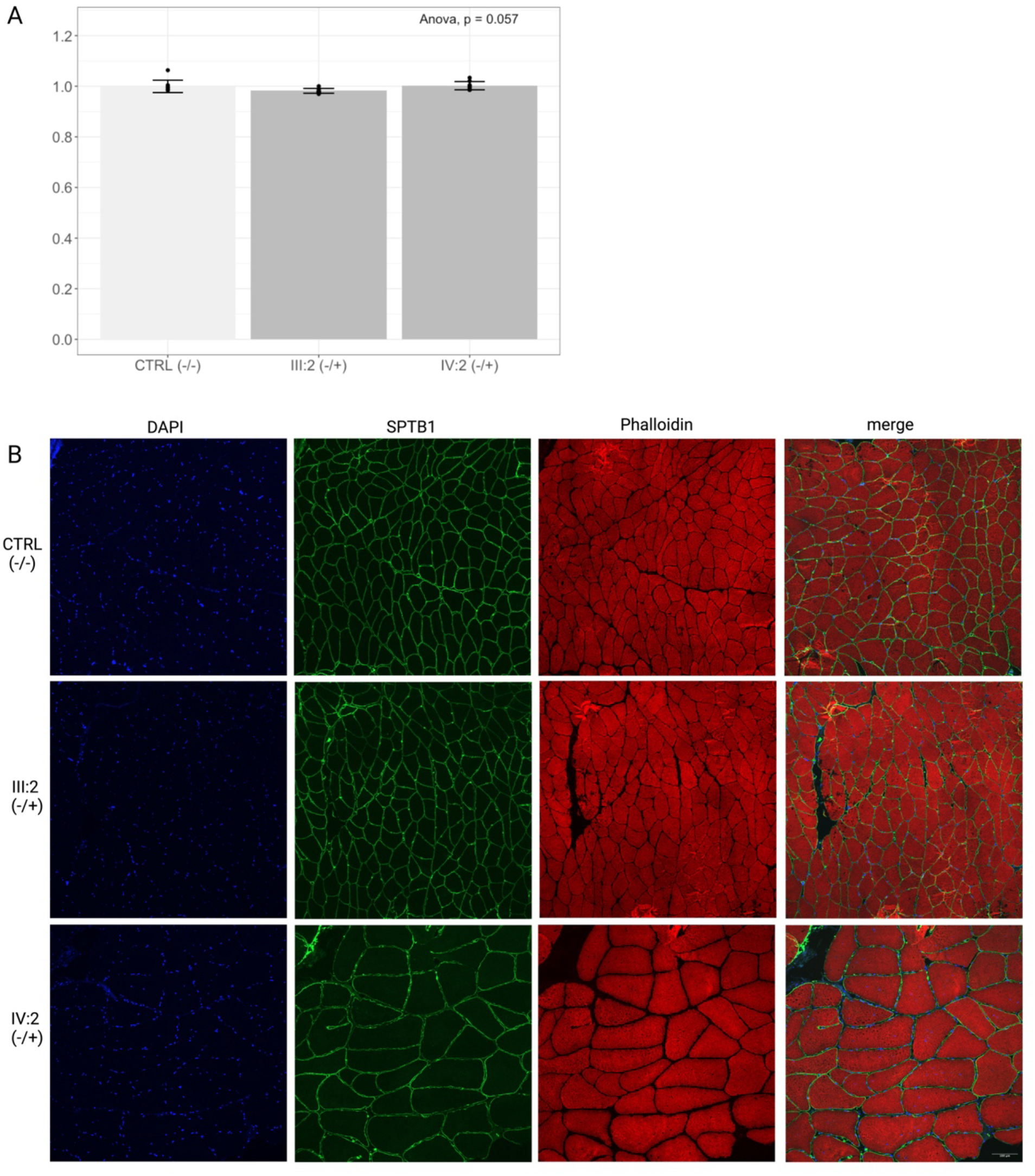
*SPTB1* muscle biopsy staining. (A) *SPTB1* intensity in ICC of muscle biopsies. (B) Representative ICC images of muscle biopsies, stained for DAPI (blue), SPTB1 (green), Phalloidin (red) and a merge image.

## References

1. Hartley T, Lemire G, Kernohan KD, Howley HE, Adams DR, Boycott KM. New Diagnostic Approaches for Undiagnosed Rare Genetic Diseases. Annu Rev Genomics Hum Genet. 2020;21:351–72.

2. Wise AL, Manolio TA, Mensah GA, Peterson JF, Roden DM, Tamburro C, et al. Genomic medicine for undiagnosed diseases. The Lancet. 2019;394(10197):533–40.

3. Zurek B, Ellwanger K, Vissers LELM, Schüle R, Synofzik M, Töpf A, et al. Solve-RD: systematic pan-European data sharing and collaborative analysis to solve rare diseases. European Journal of Human Genetics. 2021;29(9):1325–31.

4. Demidov G, Yaldiz B, Garcia-Pelaez J, de Boer E, Schuermans N, Van de Vondel L, et al. Comprehensive reanalysis for CNVs in ES data from unsolved rare disease cases results in new diagnoses. NPJ Genom Med [Internet]. 2024 Oct 26;9(1):49. Available from: https://www.nature.com/articles/s41525-024-00436-6

5. Syrbe S, Harms FL, Parrini E, Montomoli M, Mütze U, Helbig KL, et al. Delineating SPTAN1 associated phenotypes: From isolated epilepsy to encephalopathy with progressive brain atrophy. Brain. 2017 Sep 1;140(9):2322–36.

6. Beijer D, Deconinck T, de Bleecker JL, Dotti MT, Malandrini A, Andoni Urtizberea J, et al. Nonsense mutations in alpha-II spectrin in three families with juvenile onset hereditary motor neuropathy. Brain. 2019 Sep 1;142(9):2605–16.

7. De Winter J, Van de Vondel L, Ermanoska B, Monticelli A, Isapof A, Cohen E, et al. Heterozygous loss-of-function variants in SPTAN1 cause a novel early childhood onset distal myopathy with chronic neurogenic features. Florence Demurger [Internet]. 16. Available from: 10.1101/2024.09.23.24313872

8. Stankewich MC, Cianci CD, Stabach PR, Ji L, Nath A, Morrow JS. Cell organization, growth, and neural and cardiac development require αII-spectrin. J Cell Sci. 2011 Dec 1;124(23):3956–66.

9. Huang CYM, Zhang C, Zollinger DR, Leterrier C, Rasband MN. An αII spectrin-based cytoskeleton protects large-diameter myelinated axons from degeneration. Journal of Neuroscience. 2017 Nov 22;37(47):11323–34.

10. Huang CYM, Zhang C, Ho TSY, Oses-Prieto J, Burlingame AL, Lalonde J, et al. αII spectrin forms a periodic cytoskeleton at the axon initial segment and is required for nervous system function. Journal of Neuroscience. 2017;37(47):11311–22.

11. Lorenzo DN, Edwards RJ, Slavutsky AL. Spectrins: molecular organizers and targets of neurological disorders. Nature Reviews Neuroscience 2023 24:4 [Internet]. 2023 Jan 25 [cited 2023 May 15];24(4):195–212. Available from: https://www.nature.com/articles/s41583-022-00674-6

12. Laurie S. Genomic reanalysis of a Pan-European Rare Disease Resource Yields > 500 New Diagnoses.

13. Matalonga L, Hernández-Ferrer C, Piscia D, Cohen E, Cuesta I, Danis D, et al. Solving patients with rare diseases through programmatic reanalysis of genome-phenome data. European Journal of Human Genetics. 2021;29(9):1337–47.

14. Vandeweyer G, Reyniers E, Wuyts W, Rooms L, Kooy RF. CNV-WebStore : Online CNV Analysis, Storage and Interpretation. 2011;

15. Riggs ER, Andersen EF, Cherry AM, Kantarci S, Kearney H, Patel A, et al. Technical standards for the interpretation and reporting of constitutional copy-number variants: a joint consensus recommendation of the American College of Medical Genetics and Genomics (ACMG) and the Clinical Genome Resource (ClinGen). Genetics in Medicine. 2020;22(2):245–57.

16. Structural Variants in gnomAD v4 | gnomAD browser [Internet]. [cited 2024 Feb 13]. Available from: https://gnomad.broadinstitute.org/news/2023-11-v4-structural-variants/

17. Sun KY, Bai X, Chen S, Bao S, Zhang C, Kapoor M, et al. A deep catalogue of protein-coding variation in 983,578 individuals. Nature. 2024 Jul 18;631(8021):583–92.

18. Smith C, Parboosingh JS, Boycott KM, Bönnemann CG, Mah JK, Lamont RE, et al. Expansion of the GLE1-associated arthrogryposis multiplex congenita clinical spectrum. Clin Genet. 2017;91(3):426–30.

19. Kaneb HM, Folkmann AW, Belzil V V., Jao LE, Leblond CS, Girard SL, et al. Deleterious mutations in the essential mRNA metabolism factor, hGle1, in amyotrophic lateral sclerosis. Hum Mol Genet. 2015;24(5):1363–73.

20. Sert O, Ding X, Zhang C, Mi R, Hoke A, Rasband MN. The Journal of Physiology Postsynaptic β1 spectrin maintains Na + channels at the neuromuscular junction. J Physiol [Internet]. 2024;0:1–19. Available from: 10.1113/JP285894#support-information-section

21. Zhou D, Ursitti JA, Bloch RJ. Developmental expression of spectrins in rat skeletal muscle. Mol Biol Cell. 1998;9(1):47–61.

22. Duan R, Kim JH, Shilagardi K, Schiffhauer ES, Lee DM, Son S, et al. Spectrin is a mechanoresponsive protein shaping fusogenic synapse architecture during myoblast fusion. Nat Cell Biol. 2018;20(6):688–98.

